# Predicting the required pre-surgery blood volume in surgical patients based on machine learning

**DOI:** 10.1101/19008045

**Authors:** Ruilin Li, Xinyin Han, Liping Sun, Yannan Feng, Xiaolin Sun, Xiaoyu He, Yu Zhang, Haidong Zhu, Dan Zhao, Chuangchuang Dai, Zhipeng He, Shanyu Chen, Xin Wang, Weizhong Li, Xuebin Chi, Yang Yu, Beifang Niu, Deqing Wang

**Author notes:** Joint Corresponding Authors. Deqing Wang, Department of Blood Transfusion, Chinese PLA General Hospital, No. 28, Fuxing Rd, Beijing 200853, China, Beifang Niu, Computer Network Information Center, Chinese Academy of Sciences No.4 South Street, Zhongguancun, Haidian District, Beijing, China, Yang Yu, Department of Blood Transfusion, Chinese PLA General Hospital, No. 28, Fuxing Rd, Beijing 200853, China.

## Abstract

Precisely predicting the required pre-surgery blood volume (PBV) in surgical patients is a formidable challenge in China. Inaccurate estimation is associate with excessive costs, postponed surgeries and adverse outcome after surgery due to in sufficient supply or inventory. This study aimed to predict required PBV based on machine learning techniques. 181,027 medical documents over 6 years were cleaned and finally obtained 92,057 blood transfusion records. The blood transfusion and surgery related factors of perioperative patients, surgeons experience volumes and the actual volumes of transfused RBCs were extracted. 6 machine learning algorithms were used to build prediction models. The surgery patients received allogenic RBCs or without transfusion, had total volume less than 10 units, or had the latest laboratory examinations of pre-surgery within 7 days were included, providing 118,823 data points. 39 predictive factors related to the RBCs transfusion were identified. Random forest model was selected to predict the required PBV of RBCs with 72.9% accuracy and strikingly improved the accuracy by 30.4% compared with surgeons experience, where 90% of data was used for training. We tested and demonstrated that both the data-driven models and the random forest model achieved higher accuracy than surgeons experience. Furthermore, we developed a computational tool, PTRBC, to precisely estimate the required PBV in surgical patients and we believe this tool will find more applications in assisting clinician decisions, not only confined to making accurate pre-surgery blood requirement predicting.

## 1 Introduction

Insufficient supply of blood remains a challenge in many countries and this is particularly evident in China [1]. Electronic crossmatch has not been carried out yet and the vast majority of pre-surgery blood preparations are still based on serological crossmatch. The red blood cells (RBCs) transfusion is an important means to rectify anemia and improve oxygen supply. Almost 40 years age, the decision to transfuse, the “transfusion trigger,” relied on factors that affect blood transfusion, such as hemoglobin (Hb), hematocrit (Hct), fatigue, and anemia symptoms [2]. The optimal RBCs transfusion is also still a focus of controversy, and clinicians mainly rely on experience [3]. For the patients with a selective surgery, the overestimate of transfusion volume will probably bring a postponed surgery for lack of inventory and increase the cost of crossmatch, while the underestimate of transfusion volume will threaten the lives of patients because of the insufficient RBCs supply during surgery. For perioperative patients of surgical patients, an accurate PBV of RBCs plays a particularly important role both from the perspective of patients and hospitals.

The clinicians’ concern is the volume of allogeneic RBCs which are prepared by serological crossmatch prior to the surgery. Reducing unnecessary RBCs transfusion is a big trend. On the one hand, the blood supply source is insufficient [4]. On the other hand, to reduce the incidence of surgical infection and complications, it is common practice to minimize unnecessary blood transfusion. Accumulating evidence indicates that restrictive transfusion at a lower threshold of Hb concentration is safer than liberal transfusion at a higher Hb threshold [5–10]. However, restrictive transfusion could bring some adverse outcomes. For example, the mortality ratio of restrictive to liberal transfusion was 1.64 in a randomized trial [11]. The latest authoritative guidelines from the American Association of Blood Banks (AABB) showed two recommendations for hospitalized adult patients who are hemodynamically stable and neonates based on Hb level [12]. But this guideline is suitable for transfusion recommendation for hemodynamically stable patients, not for pre-surgery blood preparation of surgical patients. To get more accurate PBV of RBCs, the researches on the related pre-surgery indexes of RBC transfusion have made some progresses based on the idea of modeling. Ekhaguere et al. conducted a retrospective study of very low birth infants over 14 years, and found 13 factors affecting the RBCs transfusion [13]. However, it does not provide specific volume for the decision of clinical transfusion. Liao et. al have given quantitative scoring indicators from a clinical perspective to give the transfusion decision-making of RBCs to make the transfusion more precise [14]. But their model still in the validation phase and the blood transfusion indicators need to be further refined. Fernandes et al. performed a retrospective study on liver transplantation patients to identify pre-surgery indicators affecting blood transfusion [15], but this could not be used as a general model in clinical applications.

The development of artificial intelligence (AI) technology has brought dawn to the medical research. Machine learning is a branch of the field of AI, which application domain is extremely extensive, including almost all human cognitive fields, such as molecular biology, text processing, computer vision and robotics [16]. According to whether the model has the comprehensibility, machine learning models are divided into linear model and nonlinear model. The common linear ones include multivariate linear regression (MLR), and logistic regression (LR) [17]. The common nonlinear ones include support vector machine (SVM) [18], random forest (RF) [19], back propagation neural network (BP) [20], K nearest neighbors (KNN) [21], and XGBoost [22]. These models have been used to predict the pre-surgery transfusion volume or other predictive studies, for example, the random forest algorithm was applied to build the transfusion risk prediction model for the patients undergoing percutaneous coronary intervention [23] and regression model was applied to find the risks of the RBCs’ transfusion in obstetric patients [24]. However, the current trained models are for special types of patients or not suitable for predicting the required PBV of the RBCs in surgical patients.

In that this study, we extracted the factors directly related to RBCs transfusion from a large scale of clinical transfusion data. Based on these factors, we learned a more universal model that can be used to predict the required PBV of RBCs in surgical patients.

## 2 Materials and Methods

### 2.1 General preparation

The data were collected from electric medical records of patients who underwent surgery of the Chinese PLA General Hospital from January 1, 2011 to December 31, 2016. These records were exported from multiple real-time interaction database systems from the hospital, including HIS, LIS, PACS et al. The medical records included patients’ basic demographic information (such as admission number, name, gender, date of birth et al.), laboratory examination (including routine blood work, blood gas, biochemical indicators, blood coagulation function and routine urine analysis), blood transfusion record (such as the application volume of RBCs, the actual transfusion volume of RBCs, the storage of the RBCs et al., which covers all applications and actual transfusion information) and surgery information (such as basic surgical information, pre-surgery medication information, basic disease information et al.).

### 2.2 Data screening

Quality control for clinical data is a critical step before modeling. It determines the accuracy and reliability of subsequent results in data analysis. Moreover, it’s main goal is to identify and correct errors in medical records [25]. The review process includes three repeated cycles of screening, diagnosis, and editing [26]. Based on this idea, a detailed data screening was performed. Criteria for inclusion are below:

1. Only patients who underwent surgery were included.
2. Patients without transfusion or receiving allogenic RBCs were included, and the total RBCs transfusion volume was considered as 0 U for the former, where U represents unit(s), 1unit RBCs is derived from 200 ml whole blood, and 1.5 units RBCs means 300ml blood in China. These patients had already applied for RBCs or submitted application forms.
3. All transfusions with RBCs which occurred during the intra-surgery period were included.
4. Non-duplicate records were included.
5. Missing values were considered unavailable. The entire row of data was retained except that all values of the row were unavailable. That was, at least one record with a value was retained.
6. The latest laboratory examinations of pre-surgery within 7 days were included.
7. The same patient’s RBCs transfusion volumes were merged together when multiple transfusions were within24 hours in one surgery.
8. Patients whose total transfusion volume exceeded 10U were excluded.

The data tables were extracted and merged manually from raw discrete data tables, where 70,948 surgical no-transfusion and 92,057 surgical RBCs transfusion records were collected, respectively. Finally, 118,823 clean data points with a total of 47, 875 transfusions, 39 factors directly related to RBCs transfusion, the actual volume in patients’ and clinicians’ empirical estimates were extracted. In the subsequent analysis, the data set was divided into two groups, including patients without transfusion and receiving allogenic RBCs. The transfusion volume of RBCs varies from 1U and 10U with an interval of 0.5, where the volumes are discrete and can be divided into 20 categories.

### 2.3 Machine learning methods

Six typical and common machine learning methods were used to establish the prediction models between the required PBV of RBCs and the data features which directly related to the RBCs transfusion. The methods are LR, MLR, SVM, KNN, BP, RF and XGBoost, where LR is used to build a binary linear classification model, MLR is used to build a multi-linear regression model and the others, including SVM, KNN, BP, RF and XGBoost, are used to build nonlinear models. Their principles are briefly described below. The basic idea of SVM is to find a division hyperplane which separate samples from different categories in the sample space based on training set. SVM belongs to the discriminant model and is based on the efficient sequential minimal optimization algorithm used to solve quadratic programming problems. At present, RF has been widely used because it can compute the variable’s nonlinear effect, reflect the interaction between the variables and be not sensitive to outliers. BP is the most successful neural network learning algorithm so far. KNN is also a discriminant learning model, which is widely used because of its simple working mechanism. XGBoost is a relatively new algorithm, which is a cluster of algorithms that can elevate the weak learner to a strong learner and reduce overfitting effectively.

### 2.4 Build models

For all models, 90% of data was taken as the training set (n= 106, 940), and the remaining 10% (n=11, 883) as the test set, where the number of features was 39. To avoid the influence of imbalanced data division in the training model, the training set was generated randomly. Binary classifier means whether a patient have received RBCs (false and true), which corresponds to the decision-making of the pre-surgery preparation of blood. Multi-class classifier means the volume would be considered as classification variable, while linear regression as continuous variable. Multi-class classifier corresponds to the required PBV of RBCs for a patient. Each accuracy was derived from the maximum of the method during adjusting the model’s parameters (Table S1). Regarding logistic regression model were built and tested two models, including all the features and only the significant features as input. Then the two models were compared by analysis of variance (ANOVA) (Chi-squared test).

### 2.5 Model evaluation and validation

In this study, three groups of the volume of RBCs were acquired: the volume the patient applied for, that is clinicians’ empirical estimates (experience group), the actual transfusion volume of patients (true group) and the predicted volume generated from the models’ outputs (prediction group). The experience group and prediction group were compared to the true group, which would show the performance of the models relative to the clinicians’ empirical estimates.

The models were evaluated by the accuracy in classification or regression learning tasks, where the accuracy was calculated by the following formula[17]: 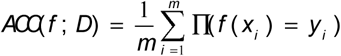. Receiver Operating Characteristic (ROC) plots were used to measure the performance of the machine learning models, where *x* = False Positive Rate (FPR) and *y* = True Positive Rate (TPR). When the ROC of a model was completely covered by another model, the latter was better. But when the two ROCs crossed, the Area Under ROC Curve (AUC) was used to measure the performance of a model. To measure linear regression, it was considered acceptable when the error was not greater than 1U according to the clinicians’ experience. The relative importance of the 39 factors were sorted according to the value of MeanDecreaseAccuracy, generated from the random forest model [19].

### 2.6 Statistical analysis

The correlation between two variables was calculated with Pearson’s correlation by IBM Statistics 22. The data report included calculations of the variables’ percentage, mean, and interquartile change. T-test and chi-square test were used to test the group difference of two groups between continuous variables and classification variables, respectively. Kruskal-Willis rank sum test was used to test the group difference of more than two groups. Psych package [27] was used for the statistical analysis. The true volume of a patient received was based on the clinicians’ experience, and the training of the prediction value was based on the true volume, so the dependent t-test was used to test the group difference of two groups, such as empirical group vs. true group and predicted group vs. true group.

## 3 Results and Discussion

### 3.1 Patient characteristics

The detailed demographic and clinical data are shown in Table 1. After the data screening process, a total of 118,823 valid RBCs transfusion records were included in this analysis, with 47,875 (40.3%) having received RBCs. A total of 165,759.5U of RBCs were transfused in 47,875 occurrences (mean, 3.46U). All factors were significantly different between subjects with and without RBCs transfusion except blood group (Table 1). In addition, the significance in different transfusion volume was also tested and the results show that 20 groups who have received RBCs are significant (p-value < 0.001) between 39 factors and the transfusion volume of RBCs.

**Table 1.**
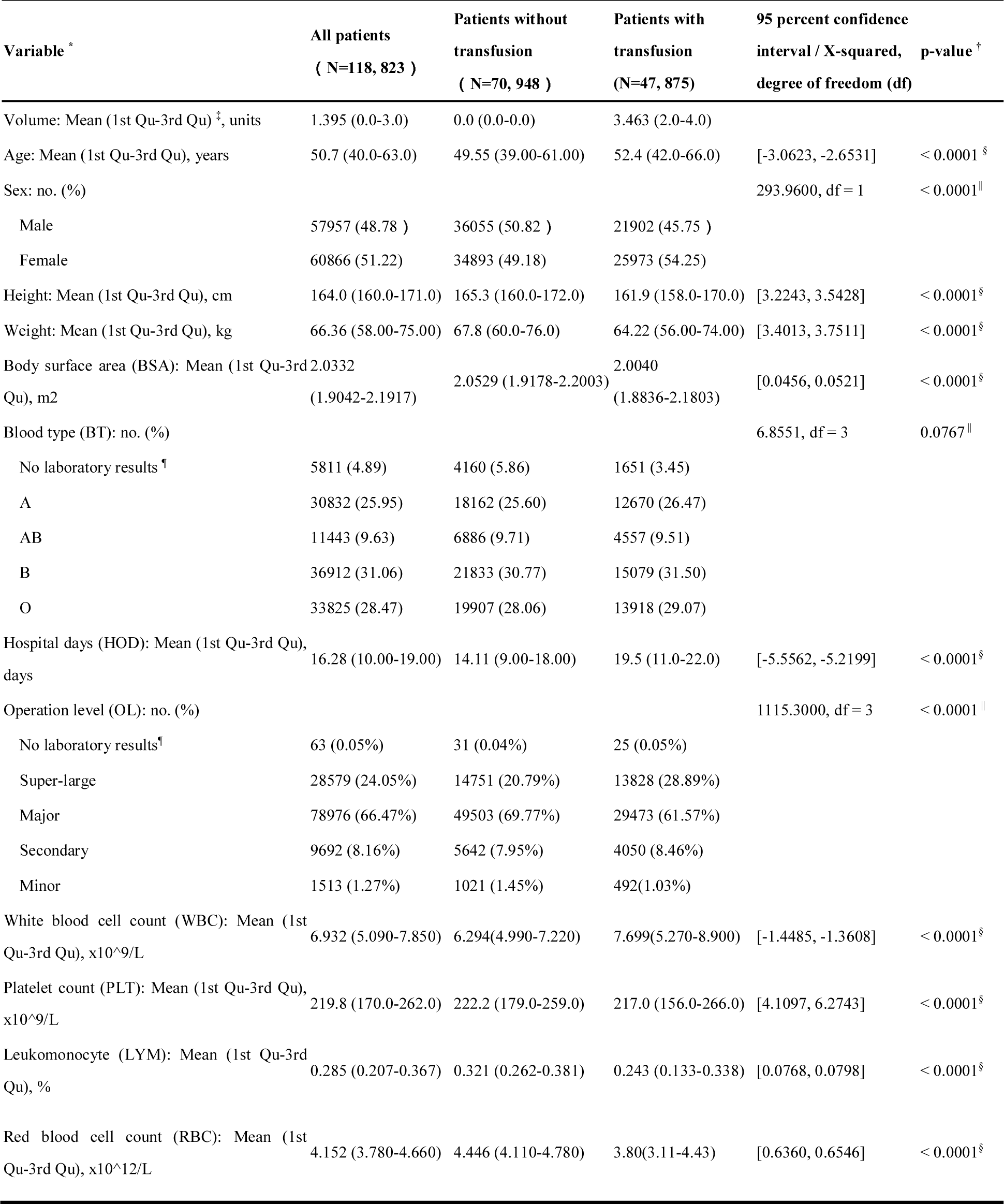
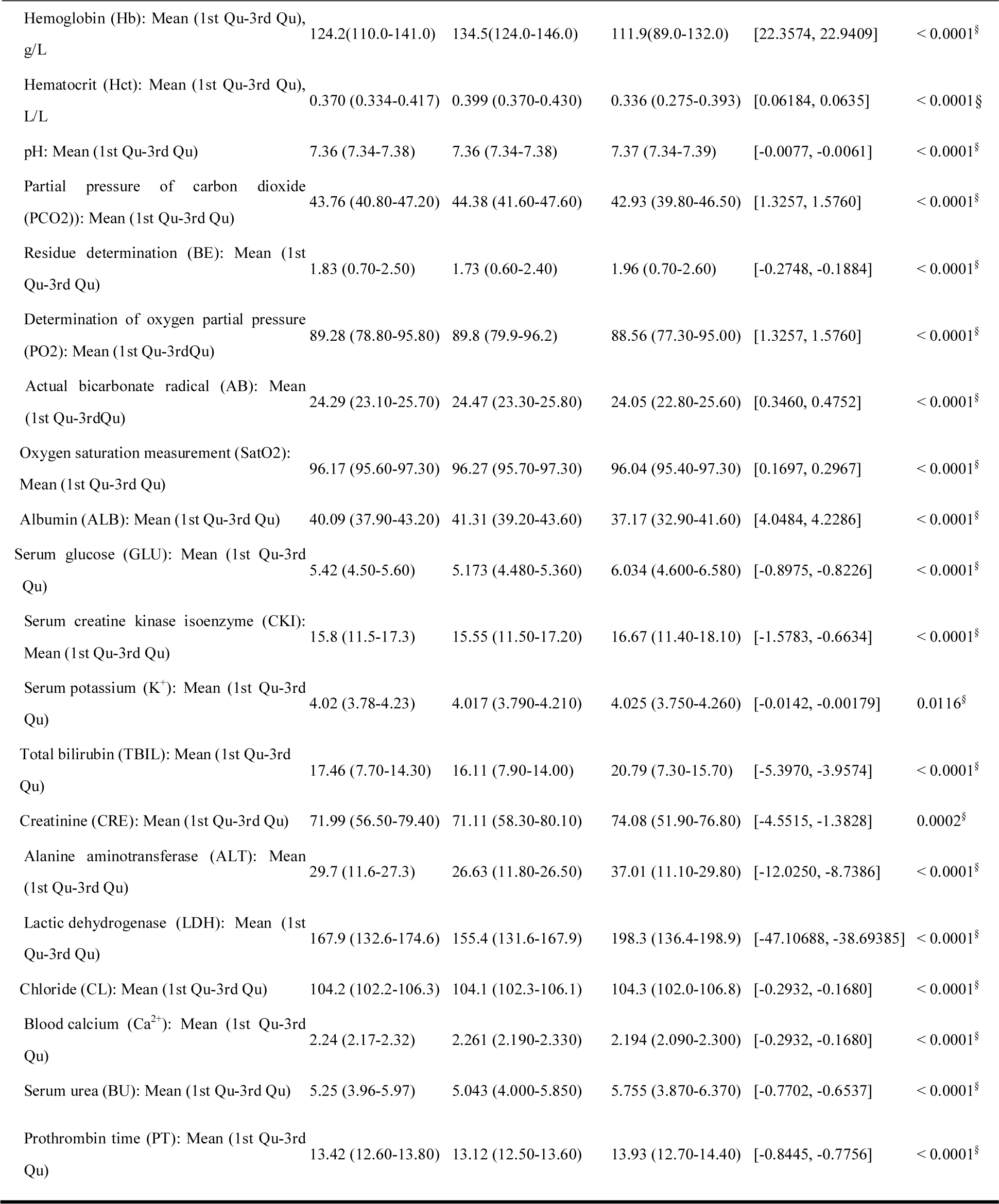
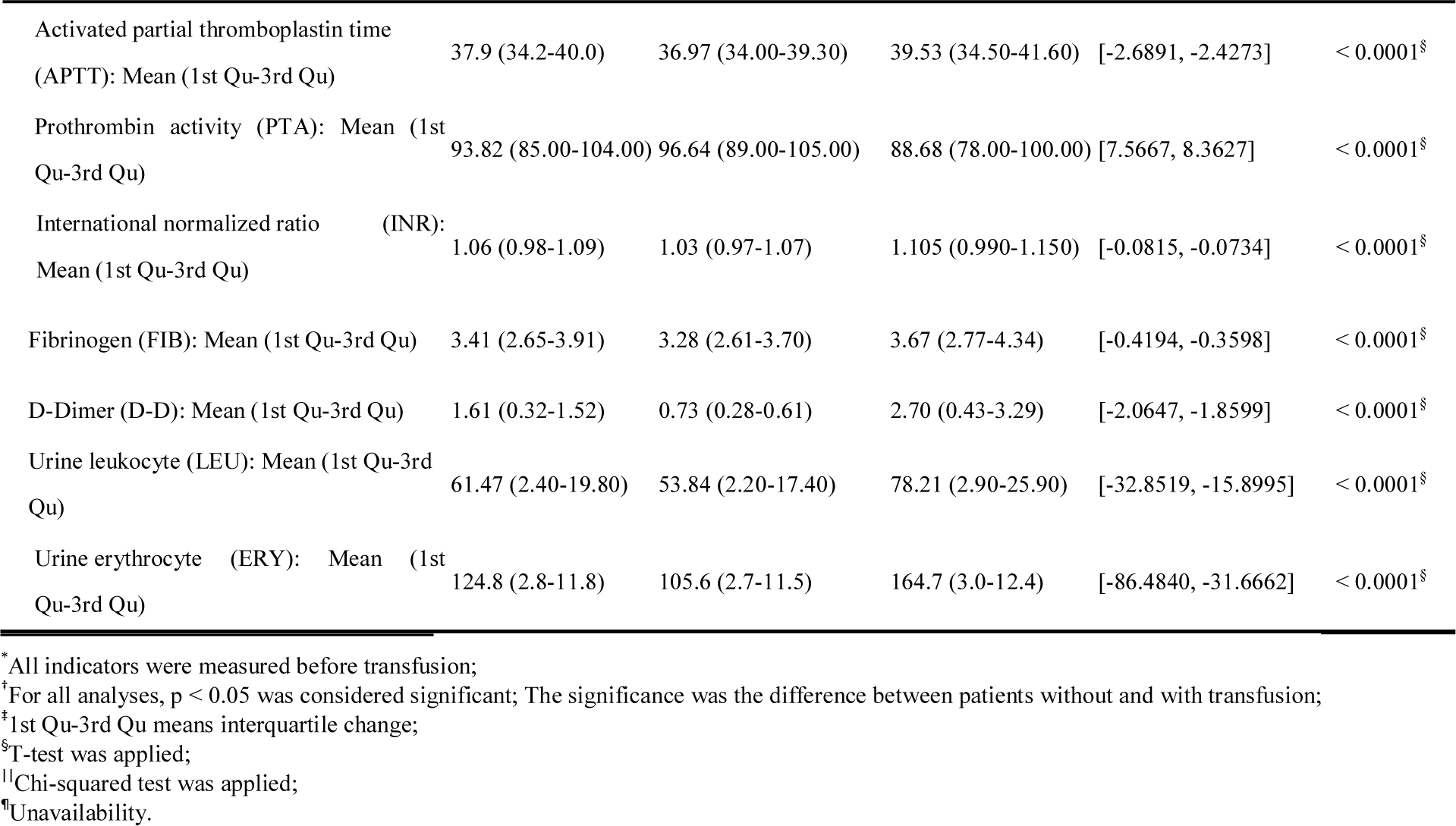
Characteristics and clinical features of the surgical patients

### 3.2 Hemoglobin concentration

The distribution of RBCs transfusion was calculated in this study. The frequency of patients who received transfusion was calculated according to pre-surgery Hb concentration, in 6 intervals: (1) <60g/L (Hb concentration < 60g/L); (2) 60g/L (60g/L ≤ Hb concentration < 70g/L); (3) 70g/L (70g/L ≤ Hb concentration < 80g/L); (4) 80g/L (80g/L ≤ Hb concentration < 90g/L); (5) 90g/L (90g/L ≤ Hb concentration ≤100g/L); (6) >100g/L (Hb concentration >100g/L). The distribution of transfusion rate in different Hb concentration intervals of pre-transfusion is shown Fig. 1. The percentages of the above 6 intervals are 0.7%, 2.8%, 7.8%, 13.7%, 12.0%, 63.0% in pre-transfusion (Fig. S1) and 0.4%, 1.0%, 2.9%, 7.6%, 16.2%, 71.6% in post-transfusion (Fig. S2).

**Fig. 1.**
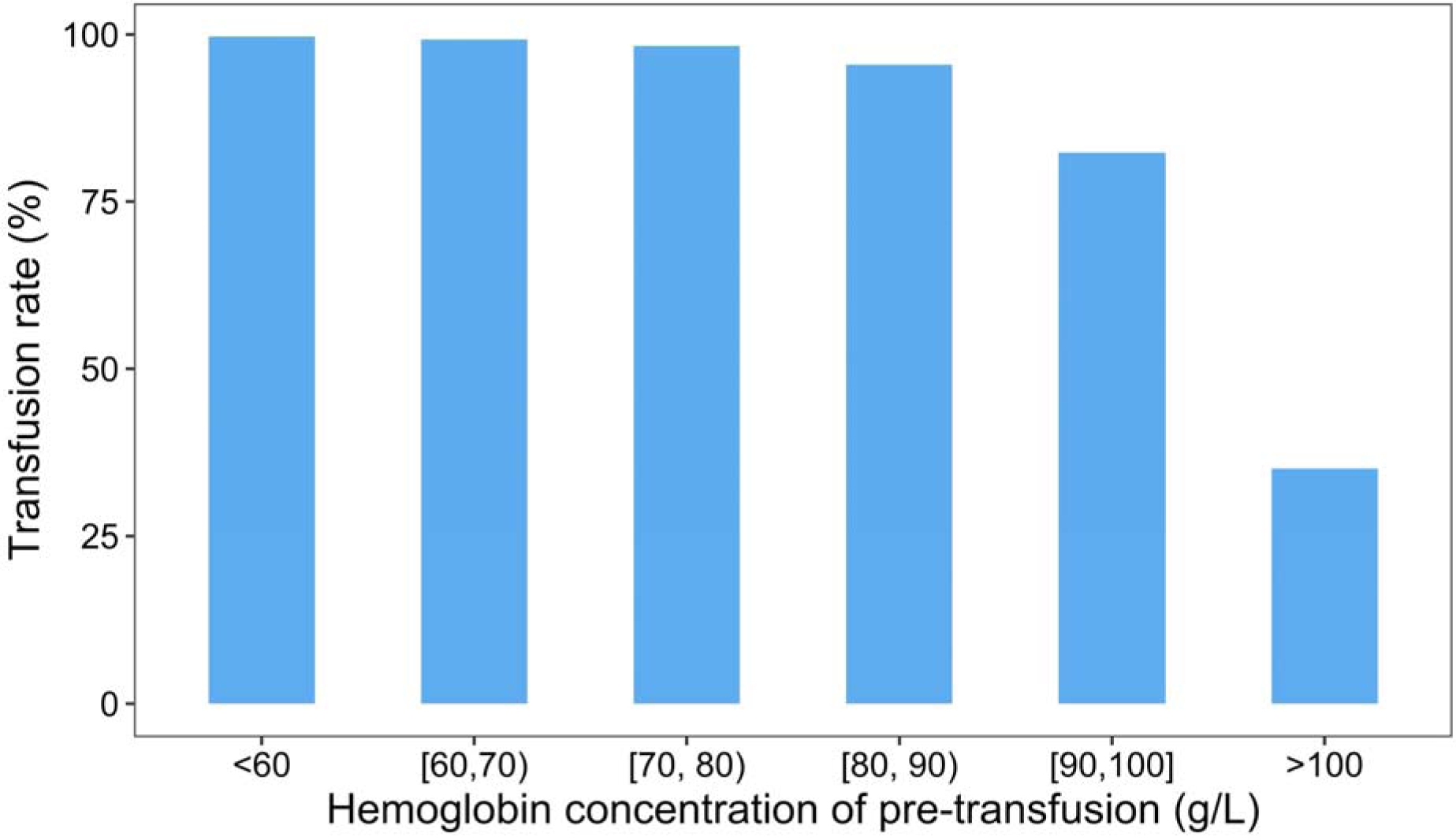
Transfusion rate in different hemoglobin concentration intervals of pre-transfusion.

### 3.3 Factors analysis

The heatmap of the correlation matrix between the 40 factors (include transfusion volume) is shown in Fig. S3. The heatmap shows the degree of relevance between features (39 clinical indicators), where 37 features are significantly correlated with one or more other features except for urine leukocyte (LEU) and urine erythrocyte (ERY) (Pearson correlation when the confidence was 0.05). We also analyzed the effect of storage of RBCs, and the result showed that the storage time of RBCs had a significant positive influence on the volume of RBCs transfusion (Pearson two-tailed test, correlation coefficients: 0.071, p-value < 0.01).

### 3.4 Modeling and results

Data from this study, were used to test the performance of models generated from 6 typical machine learning algorithms. Two methods of predicting of each machine learning algorithms were binary classifier and multi-classifier. The comparison of accuracies between clinicians’ experience recommendation and machine learning models are shown in Fig. 2. All the machine learning models’ performance was superior clinicians’ experience recommendation (experience group) both in binary and multi-class classifier. The results of two LR models including all the features and only the significant features as input showed that the fitting degree of the two models was the same under different predictive variables (P=0.2888>0.05), so the prediction can be preformed with only the significant features as input (RBC, PCO2, SatO2, CKI, K^+^, Ca^2+^, INR and LRU were excluded because their regression coefficients were not significant). Regarding MLR model, 4 features including GLU, CKI, ALT and INR were excluded because they would reduce the model’s quality in the process of model optimization.

**Fig. 2.**
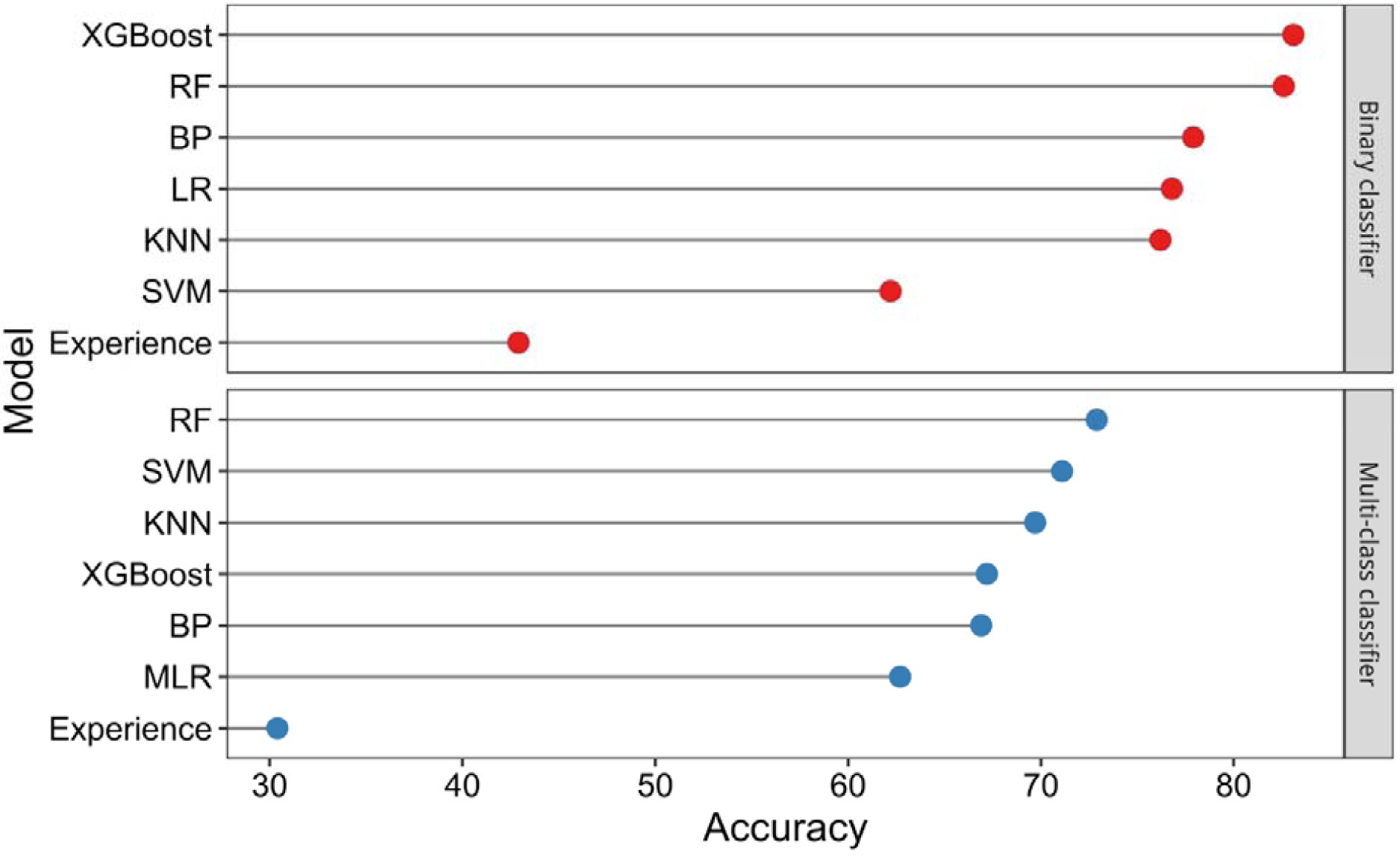
Comparison of accuracies between clinicians’ experience recommendation and models. In multi-class classifier group, the accuracies are normalized values in regression. “Experience” represents the clinician’s recommendation.

The experience group and prediction group were compared to the true group. Using the dependent T-test, the result showed that True vs. Experience, 95% CI: [1.7807, 1.8818], p-value < 0.001, the mean of the differences is 1.8313; True vs. Prediction, 95% CI: [−0.8960, −0.8296], p-value < 0.001, the mean of the differences is −0.8628 (Fig. 3). The result showed that the mean of prediction group was much closer to the true group.

**Fig. 3.**
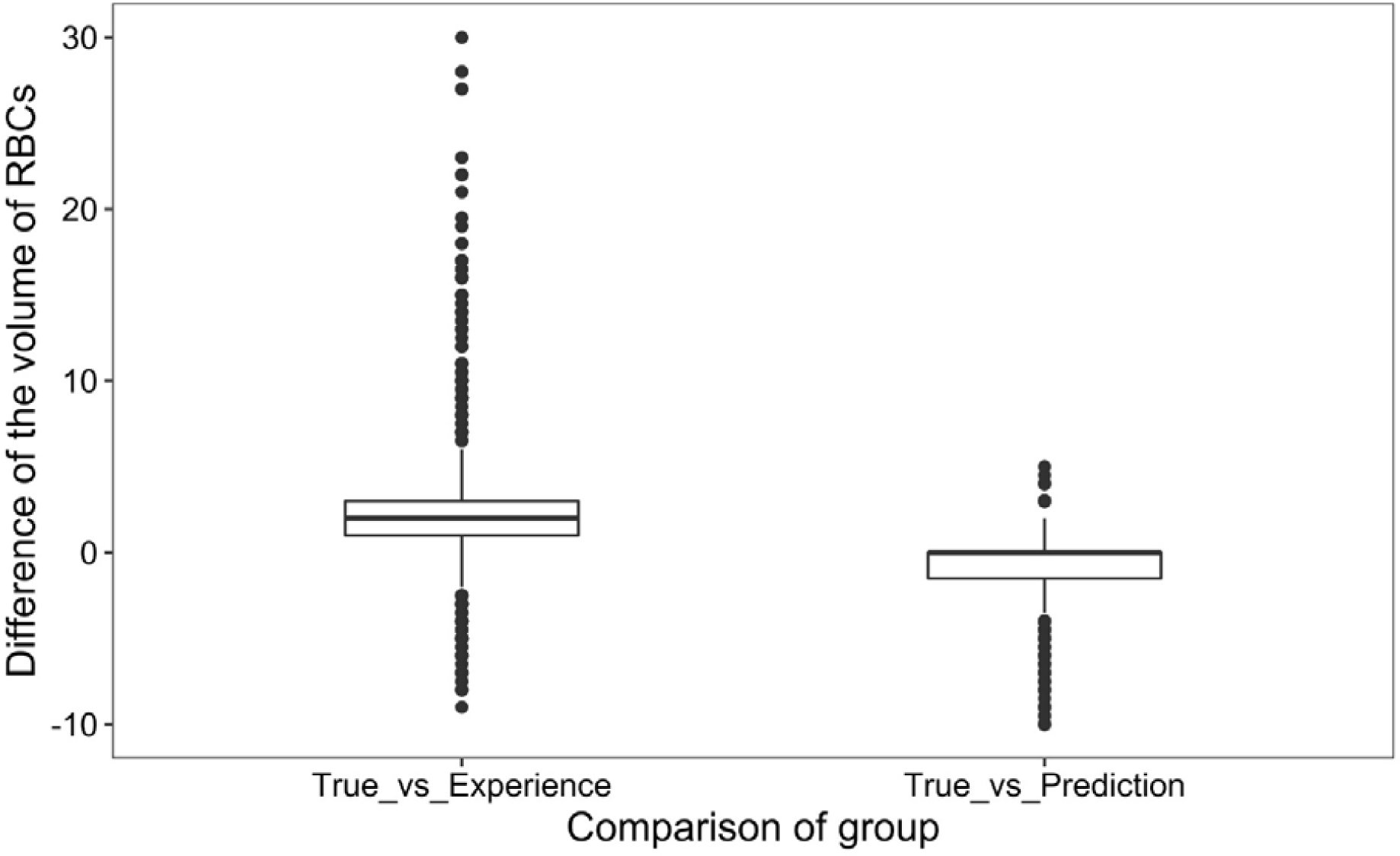
Comparison of the difference of the volume RBCs between true vs. experience and prediction.

According to the results of Fig. 2, RF model obtained the highest accuracy in predicting the required PBV of RBCs under the condition of multi-class classifier. Based on this optimal model, the individual categories’ evaluation was performed and features’ importance were ranked. The ROC curve of the Random Forest model is shown in Fig. 4a. The accuracy of each class changed from 82.31% to 99.97% (Fig. 4b), which indicated that the accuracy of each category was more than 82%. The indicators considered clinically important are included in the top 20 factors for the RBCs transfusion, like age, OL, weight, Hct, Hb, RBC and height, LYM, BSA and HOD (Fig. 4c).

**Fig. 4.**
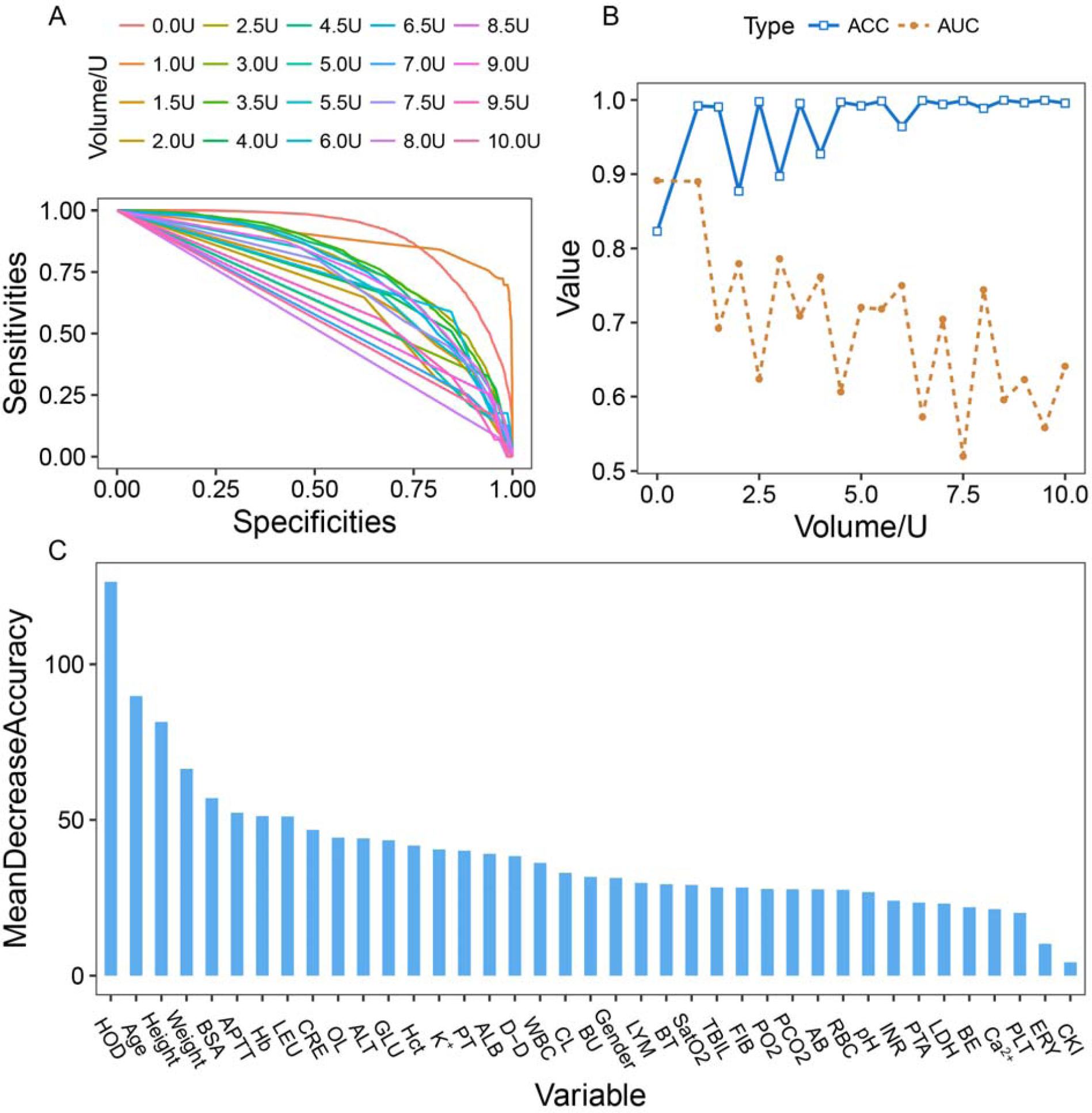
**a** Receiver Operating Characteristic (ROC) curve of each class in the Random Forest model. **b** Comparison of area under the curve (AUC) and accuracy (ACC) in each class. **c** Importance of factors in the Random Forest model. MeanDecreaseAccuracy means the reduction in the degree of accuracy when the value of a variable is changed into a random number. The larger the value is, the greater the importance of the variable.

### 3.5 Implementation and availability

We developed a lightweight prediction tool, named as PTRBC (Prediction of Transfusion with Red Blood Cells). PTRBC was implemented in Perl and R programming. It is freely available for non-commercial use at https://github.com/niu-lab/PTRBC. We also used the R packages from the Comprehensive R Archive Network (https://cran.r-project.org/) including DMwR, randmoForest, xgboost, e1071, gmodels and class.

### 3.6 Discussion

Clinical data mining is an important trend in the development of modern medicine in the information age. The common machine learning solutions include classification, clustering, prediction, and regression [28]. Machine learning is widely used in the field of medicine and has achieved good results, such as the automatic sensing method was developed to sensitively detect unstained malaria-infected RBCs [29]. Machine learning acquires the clinical experience through the means of calculating, although the experience is not entirely correct, the prediction will tend to be more accurate as the data is constantly updated and the model is repeatedly trained. Therefore, to build data-driven decision-making models can help to improve the quality of medical. Before the implementation of the electronic crossmatch, the accurate prediction of the pre-surgical preparation of blood can be realized via calculation, which will effectively save blood consumption, improve the accuracy of blood supply and relieve the insufficient supply of blood challenge.

In clinical blood transfusion, surgeons often focus on two issues: whether to give the RBCs transfusion and how many a patient should be given. The formula for calculation of blood transfusion volume based on weight and Hb concentration had been extensively studied, specially, for the critically ill patients [30] and children [31]. But these models are for specific groups of patients. Based on the above issues, we have built six models based on the overall clinical context and alternative therapies besides the Hb concentration to predict the transfusion decision and transfusion volume. Our study showed that there were 37 significant factors between the two groups as shown in the last column of Table 1. We found that the rate blood transfusion with RBCs decreased with the increase of hemoglobin concentration (Fig. 1). For patients with transfusion, more than 63.0% of patients received a RBCs transfusion in our cohort, although their Hb concentration of pre-transfusion exceeded 100 g/L (Fig. S1). However, the individualized blood transfusion should be considered for these patients whose Hb concentration of pre-transfusion do not meet the guidelines. The percentage of post-transfusion Hb concentration more than 9g/L is 87.9% (Fig. S2), which means that most patients achieved the recommended Hb concentration in AABB guidelines (9 g/L).

The factors affecting blood transfusion are more complicated, including the patient’s own characteristics and the characteristic of the RBCs to be transfused. Specifically, the patient’s own factors include multiple blood transfusions, immune system diseases, fatigue, dyspnea, mechanical ventilation, low venous oxygen saturation (SvO_2_), blood flow loss, Hb, Hct, inadequate oxygen supply, blood pressure, heart rate, body temperature, and so on [32]. The storage time of blood is also a significant factor [33], and our results show that storage time is also a significant factor. But the storage time of RBCs was not used when building models because our cohort included the patients given more than one bagged RBCs in different storage time in a blood transfusion. In the current report, 39 factors related to RBCs transfusion have been analyzed, and the correlations showed that 37 factors are significant (p-value < 0.05) (Table 1). But in the process of modeling, all 39 factors were all considered in our models. Because the accuracy would decrease with the decrease of the number of features for the nonlinear machine learning models. Therefore, this study took all features as input for nonlinear machine learning models.

The results showed that random forest and XGBoost models were optimal in predicting the volume of RBCs transfusion in all the training models. The XGBoost model can be used to the assisted decision-making in pre-surgical preparation volume of RBCs (reached 83.1% accuracy), and the random forest model can be used to given the preparation volume of RBCs (reached 72.9% accuracy) (Fig. 2). Nevertheless, the volume of clinicians’ empirical estimates only up to 42.9% and 30.4% accuracy, respectively. We observed that the mean value in prediction group was closer to the real group than that in the empirical group (−0.8628 vs. 1.8313) (Fig. 3). The mean value of prediction group was lower than the real group, because there were too many samples who were no-transfusion in the training set, which may lead to some model deviation. Overall, the prediction group had minor fluctuation. Previous studies have been based on Hb and weight to calculate the expected volume of RBCs [30] and the top 10 important factors (Fig. 4c) show that these two indicators are high on the list, where the important factors are consistent with the previous results.

Our model has achieved good results, but there are some minor limitations. Clinically speaking, the model in this study was not applicable for patients who underwent more than 10U RBCs transfusion to minimize the introduce of disturbance. From a modeling perspective, hospital days, though it is a post-surgical factor, was included in our model because it had a high weight on the model.

## 4 Conclusions

Existing models associated with the required PBV predicting have many shortcomings: not suitable for prediction of pre-surgical RBCs preparation, without generality, and fewer factors were considered. Meanwhile, clinicians experience that used in everyday practice result in lower accuracy, compared with the actual transfusion volume of patients. In this paper, we presented a model combination method and this method can achieve higher accuracy than clinicians experience, not only in pre-surgical blood requirement decision but also in the required volume of allogeneic RBCs. Moreover, a new prediction tool, PTRBC, is developed in this paper, and this tool can be used to give the specific required PBV of RBCs. We believe this to be a prospective study predicting the required PBV in surgical patients for the countries who faced with the dilemma of electronic crossmatch, which will provide a novel method for studying other blood components, such as plasma or platelet.

## Data Availability

If you want to use the data in this article, please contact the corresponding author for permission.

## Acknowledgements

This work was supported by the National Natural Science Foundation of China (Grant No. 31771466), the National Key R&D Program of China (Grant nos. 2016YFC0503607, 2018YFB0203903), the Special Project of Informatization of Chinese Academy of Sciences of China (Grant No. XXH13504-08), the Strategic Pilot Science and Technology Project of Chinese Academy of Sciences (Grant No. XDA12010000), the Key Project-subtopic of “13th five-year plan” Military Logistics Research (Grant No. BWS16J006), the Medical Big Data R&D Project of PLA General Hospital (Grant No. 2016MBD-023).

## Authors’Contribution

This study was designed by D. W., Y. Y. and B. N. R. L. performed the research and drafted the manuscript. X. H. drew all the figures. L. S, Y. F., X. S., Z. D. and X. W. provided the methods and clinical guidance. H. Z. and Y. Z. contributed the code debugging. W. L., X. Y. H., C. D, Z. H, S. C., and X. C. edited and revised the manuscript.

## Compliance with Ethical Standards

The study protocol was approved by the Ethics Committee of Chinese PLA General Hospital, in accordance with the Declaration of Helsinki.

